# Persistence, prevalence, and polymorphism of sequelae after COVID-19 in young adults

**DOI:** 10.1101/2022.02.11.22270836

**Authors:** Jeremy Werner Deuel, Elisa Lauria, Thibault Lovey, Sandrine Zweifel, Mara Isabella Meier, Roland Züst, Nejla Gültekin, Andreas Stettbacher, Patricia Schlagenhauf

## Abstract

**Background:** COVID-19 sequelae are poorly defined with an ambiguous pathophysiology. Persistent sequelae could have global, public health and societal ramifications. We aimed to describe sequelae presenting more than six months after COVID-19 in non-hospitalized young adults.

**Methods:** A prospective, longitudinal cohort study followed-up on young Swiss Armed Forces (SAF) personnel. The comprehensive test battery was administered during a single full day of testing at the University of Zürich. It quantified the impact of SARS-CoV-2 infection on cardiovascular, pulmonary, neurological, renal, ophthalmological, male reproductive, psychological, and general health in addition to laboratory parameters.

**Results:** We included 501 participants (5.6% females) with a median age of 21 years (range 19-29). Cases of previous COVID -19 (>6 months (mean 10 months) since diagnosis, n=177) were compared with never infected controls (n=248). We also included more recent COVID-19 cases (≤6 months, n=19) and asymptomatically infected individuals (n=49). We found a significant trend towards metabolic disorders, higher Body Mass Index (BMI) (p=0.03), lower aerobic threshold (p=0.007), higher blood cholesterol (p<0.001) and low-density lipoprotein LDL levels (p<0.001) in participants> 6 months post Covid-19 when compared to controls. There were no significant differences in psychosocial questionnaire scores, ophthalmological outcomes, sperm quality or motility between controls and those infected more than 6 months previously with SARS-CoV-2.

**Conclusions:** Young, previously healthy, individuals largely recover from mild infection and the multi-system impact of the infection is less that seen in older or hospitalized patients. These results may be extrapolated to health-care workers and other young workforce adults. However, the constellation of higher body mass index, dyslipidemia and lower physical endurance 6 months post COVID-19 is suggestive of a higher risk of developing metabolic disorders and possible cardiovascular complications. These findings will guide investigation and follow-up management.

## Introduction

The COVID-19 pandemic, caused by the coronavirus SARS-CoV-2, is ongoing, with intense global transmission. As of January 24^th^, 2022, more than 356 million persons have been infected (1). Mounting evidence indicates that SARS-CoV-2 is a multisystem infection. Data on how long symptoms persist and the intermediate- and long-term sequelae of the infections have scarcely been researched. Available original research tends to focus on patients who have been hospitalized (2,3) or restricts evaluations to a single organ system (3). Studies to date show that persisting sequelae of COVID-19 disease are common in persons with risk factors: older adults, smokers, and those with underlying comorbidities such as hypertension, obesity, diabetes, cardiovascular disease, chronic lung disease, chronic kidney disease, chronic liver disease, cerebrovascular disease, cancer and immunodeficiency. Sequelae of infection have however also been observed following milder SARS-CoV-2 infections (4, 5) in population-based studies that followed up on prescription data or in electronic health databases (6) or on patients presenting to post COVID clinics (5). In a telephone survey (7) in adults who tested positive for SARS-CoV-2, 35% of 274 symptomatic respondents, including 26% amongst those aged 18-34 years, reported not having returned to their usual state of health two weeks or more after testing. The Pan American Health Organization PAHO has issued an epidemiological alert on the need for information regarding the complications and sequelae of COVID-19 (8). The World Health Organization (WHO) has added “*post COVID-19 condition*” to the International Classification of Diseases codes to describe a condition that occurs people following probable or confirmed SARS-CoV-2 infection with symptoms that last for at least two months and that cannot be explained by an alternative diagnosis (9). In late 2020, a Long COVID Forum brought together sufferers, stakeholders, researchers, and policy makers including the WHO to identify research gaps and a core recommendation here was to expand research beyond hospitalized patients (10). Systematic evaluation of multi-organ function using sensitive test batteries with quantitative outcomes and matched negative controls are clearly needed for discrete population groups. Such data are particularly important in the context of young adults who constitute a large proportion of any country’s health care workers and/or other workforce members.

We aimed to design a minimally invasive test battery that could comprehensively evaluate and follow-up on longer-term sequelae of SARS-CoV-2 infection with a focus on pulmonary, cardiovascular, neurological, renal, ophthalmological, male reproductive, psychological, and general health in addition to serological and laboratory parameters. The battery components were based on the results of our systematic review of COVID-19 sequelae in young previously healthy adults (11). The goal of this LoCoMo (**Lo**ng **CO**VID in **M**ilitary **O**rganisations) study was to quantitatively assess the impact of infection on multi-organ systems in a cohort of young, healthy, mainly male, Swiss Army recruits. The Swiss Armed Forces (SAF) has a conscription system with a 10-year duration of mandatory service. The soldiers, who are recruited as young adults aged between 18 and 30 years, return annually for repetition courses. This LoCoMo study follows up on recruits who tested either positive or negative at Swiss Army bases in 2020-2021.

## Methods

### Study Design, Setting and Recruitment

LoCoMo is a prospective, longitudinal cohort study approved by the Swiss Zürich Cantonal Ethics Committee (BASEC-Nr. 2021-00256). Registration https://clinicaltrials.gov/ct2/show/NCT04942249. Potential participants aged 18-30, with recent military service in 2020-2021, received a written invitation to voluntarily enroll in the LoCoMo study using an online booking tool. The volunteers took part in one day of intensive testing (Figure 1) at the University of Zürich and associated clinics at the University Hospital of Zürich. After check-in and a saliva SARS-CoV-2 rapid antigen test (COVID-19 Antigen Detection Kit (Colloidal Gold) Zhuhai Lituo Biotechnology Co., Ltd) to exclude concurrent SARS-CoV-2 infection, participants completed a consent form and answered simple baseline questions (age, sex, body height, body weight, test status, co-morbidities, smoker status, educational level, vaccination status). All questionnaires, consent forms and flyers were available in German, French and Italian languages. The volunteers underwent all the tests outlined in the test battery and provided venous blood and saliva for processing in the Hematology and Clinical Chemistry divisions, at the in-house lab and biobank and at the Spiez Laboratory. All procedures in the test battery were done by specially trained scientists and physicians. A sperm count was optional, (Figure 1). The additional inclusion criteria here were male sex and no known reproductive anomality. Several questionnaires were self-administered on iPads that were available to the participants throughout the day. Assistants were available to bring participants to their allotted appointments in the Ophthalmology and Andrology clinics. At the end of the testing day, participants did a “check-out” to ensure that all test-components and questionnaires had been completed. All questionnaire answers and data on the study participants were stored in the REDCap secure database with an “auto-archiver”. Completed files were stored in a secure file Repository.

**Figure 1:**
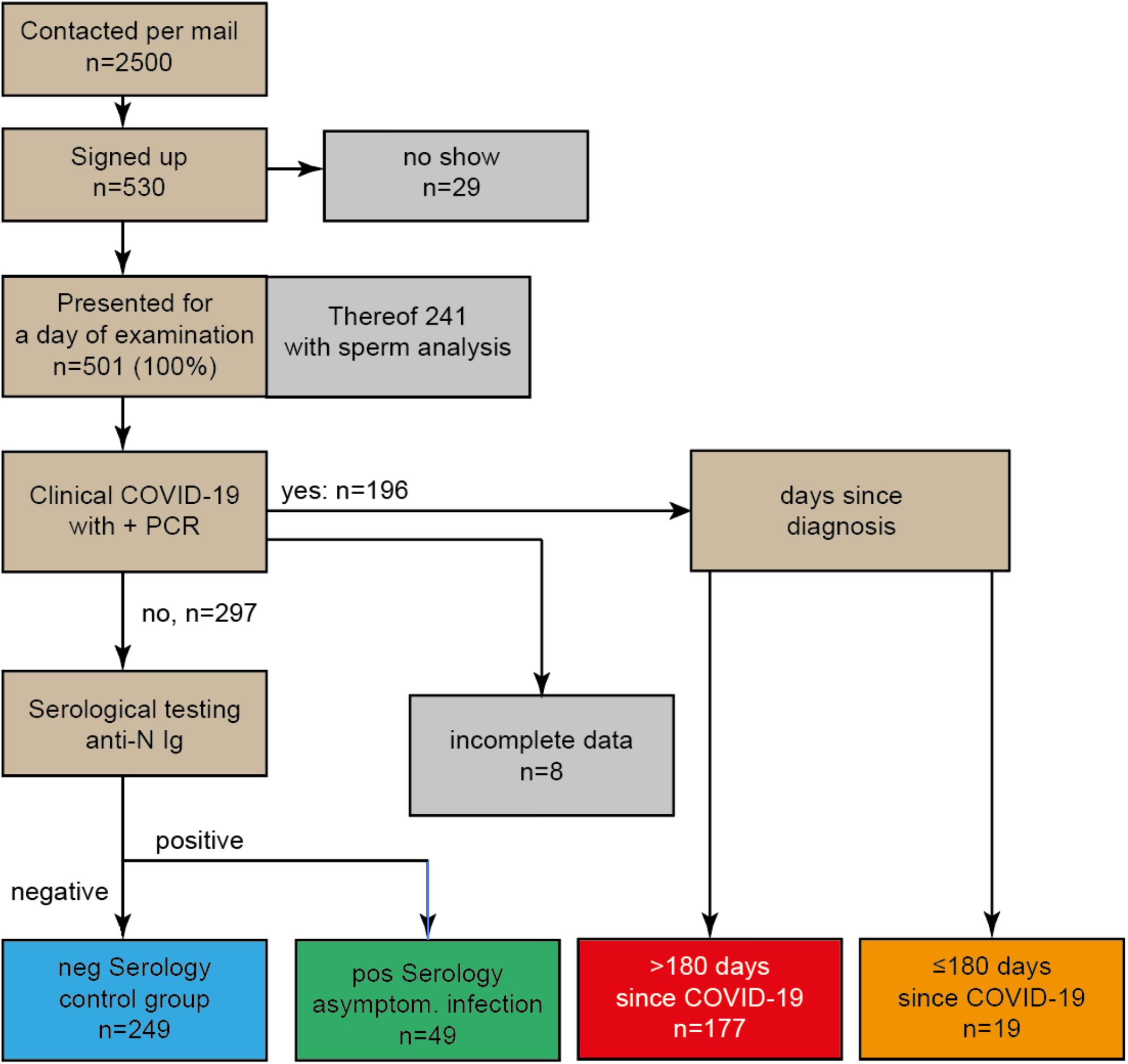
Flow chart of the study. 2500 members of the Swiss Armed Forces were contacted by mail, 530 thereof (21%) agreed to participate in the study and 501 (95%) presented for a full day of testing in Zurich. Participants were grouped according to their COVID-19 status into a control group (no clinical or serological evidence of past Infection with SARS-CoV-2), an asymptomatic group (no clinical evidence but positive serology), and patients after confirmed COVID-19. The latter were further sub grouped according to the duration since the day of diagnosis of COVID-19 into recent and non-recent COVID-19.

### Test Battery

Based on our earlier systematic literature review of possible COVID-19 sequelae in young persons, a non-invasive test battery evaluated the following:

#### General symptoms

Fatigue was assessed using the validated Chalder Fatigue Scale (CFQ-11) and also using the Profile of Moods States 2 (POMS2). Kidney function was assessed using the estimated glomerular filtration rate (eGFR) and creatinine and cystatin C levels. Blood sampling allowed for measurement of routine laboratory parameters including white cell counts (counts and full differential) and C-Reactive Protein (CRP). Serum and saliva were bio-banked.

#### Pulmonary/Respiratory System

Lung function was assessed using spirometry (12). CO diffusion capacity testing was performed to provide an index of damage to microcirculation or interstitial damage and to estimate the total lung capacity (TLC). Expiratory NO (FE-NO) assessment provided an indication of inflammatory processes within the lung.

#### Cardiovascular sequelae

We measured N-terminal pro-Brain natriuretic peptide (NT-proBNP) as a marker for congestive heart failure as well as Troponin T to test for myocarditis.

#### CPET

A cardio-pulmonary exercise test (CPET) was performed on a treadmill while measuring work rate, heart rate, blood pressure, in- and expiratory CO_2_ and O_2_ concentration as well as gas flow and analysed as previously described elsewhere (13)

#### Chemosensory

Olfactory function was assessed using the “Sniffin’ Sticks” test (14), providing a quantitative outcome called the composite “Threshold-Discrimination-Identification” (TDI) score, that indicates normosmia (TDI ≥31), hyposmia (TDI <31), or functional anosmia (TDI ≤ 16). Gustatory performance was measured using “Taste Strips”, (15) in a test to investigate the ability to perceive four primary tastes (sweet, salty, sour and bitter). Quantitative scores provided the follow assessments: normal, mild hypogeusia, moderate hypogeusia, severe hypogeusia, ageusia.

#### Ophthalmological sequelae

All subjects underwent a complete ophthalmic examination including optical coherence tomography angiography (OCTA) scanning, color fundus photography (CF), ultra-wide field (UWF) CF and autofluorescence imaging (16).

#### Psychological sequelae, emotional health

The following questionnaires were self-administered: *Quality of Life -EQ-5D-5L, COVID-19-PTSD, Zung Self-rating Depression Scale (ZSDS), Beck’s II Depression Scale, State-Trait Anxiety Inventory form-Y (STAI-Y*), and *Profile of Mood States 2* **Male fertility:** A standard WHO sperm count was performed to evaluate semen volume, sperm concentration, motility and morphology (17). Male sex hormones were measured.

#### Neutralizing Antibodies

Neutralizing antibody (nAb) titers of vaccinated, recovered, and recovered/vaccinated, and control groups were evaluated using a previously established methodology (18).

### Statistical Analysis

Data were analysed using R statistical Software Version 4.1.2, R Foundation for statistical computing, Vienna, Austria (19). For each outcome or test result, (Welch Two Sample t-test, Wilcox test) we used Odds Ratios to compare post COVID-19 volunteers to SARS-CoV-2 negative individuals by fitting a generalized linear model and calculating odds ratios from there using the package “oddsratio” Version 1.0.2. Graphics were generated by ggplot2. Subgroup analyses were performed to evaluate differences based on interval elapsed since infection (<6 months versus > 6 months) and the severity of infections (asymptomatic versus symptomatic).

### Role of the funding source

This study was funded by the Swiss Armed Forces (SAF). The initial protocol was evaluated by the Army Research Committee for input regarding study duration, costs and outcome measures. NG and AS were instrumental in initiating the study, in finances and in liaising at the army/university interface to ensure incentives such as military service days. NG and AS contributed to the revisions of the final paper and to the approval of the final manuscript. The funding source had no access to the data or role in the analyses.

## Results

The baseline characteristics of participants are shown in Table 1. We included 501 participants (5.6% females) with a median age of 21 years (range 19-29). Cases of previous COVID-19 (>180 days since diagnosis, n=177) were compared with never infected controls (n=248) (Figure 1). We also included recent COVID-19 cases (≤180 days, n=19) and asymptomatically infected individuals (n=49, with serological evidence of infection but without confirmed COVID-19 (Figure 1). We found a significant trend towards a constellation of metabolic syndrome, with higher Body Mass Index (BMI) (p=0.03), lower aerobic threshold (p=0.007), higher blood cholesterol (p<0.001) and LDL levels (p<0.001) in participants more than 6 months post COVID-19 when compared to controls (Figure 2, Figure 3). Participants in the more than 6-months post COVID-19 group reported more “fatigue” on the POMS scale compared to asymptomatic SARS-CoV-2 infected (p=0.0005). Otherwise, there were no significant differences in psychosocial questionnaire scores, sperm quality or motility between controls and those infected more than 6 months previously with SARS-CoV-2 (Appendix 1). In a subgroup analysis comparing recent (<6 months since diagnosis) COVID-19 cases versus less recent (>6 months since diagnosis) (Figure 4) significant hyposomia (TDI <31) (p=0.027) was observed in the more recent cases but not in less recent cases (Appendix 1). The andrology results showed significantly (p=0.004) poorer motile sperm count in participants with recent COVID-19 compared to controls and less recent COVID-19 (p=0.03), although no difference was observed more than 180d after COVID-19. STAI S scores of anxiety levels were significantly higher in recent COVID-19 compared to controls (p=0.022) or to less recent infections (p=0.031). PTSD-19 scores and Beck’s Depression scale showed higher psychological burdens in the recent COVID-19 group compared to less recent COVID-19 cases.

**Table 1:** Demographics of all groups evaluated

| **Demographics** | **Control** | **Post COVID-19** | **Recent COVID** | **Asymptomatic Infection** |
| --- | --- | --- | --- | --- |
| Number of participants | 249 | 177 | 19 | 46 |
| Age (years) | 21 (21-21) | 22 (21-24) | 21 (21-22) | 21 (20-22) |
| Days since COVID-19 | - | 317 (272-414) | 101 (83-155) | - |
| Anti-N (COBAS, A.U.) | 0.064 (0.059-0.067) | 3.61 (1.00-10.0) | 14.4 (4.52-39.3) | 4.62 (2.06-8.44) |
| Vaccinated* | 24.1% | 33.3% | 47.4% | 34.8% |
| Males | 94.8% | 93.8% | 94.7% | 91.3% |

Median, 25/75% quantile in brackets, \*one or more times

**Figure 2:**
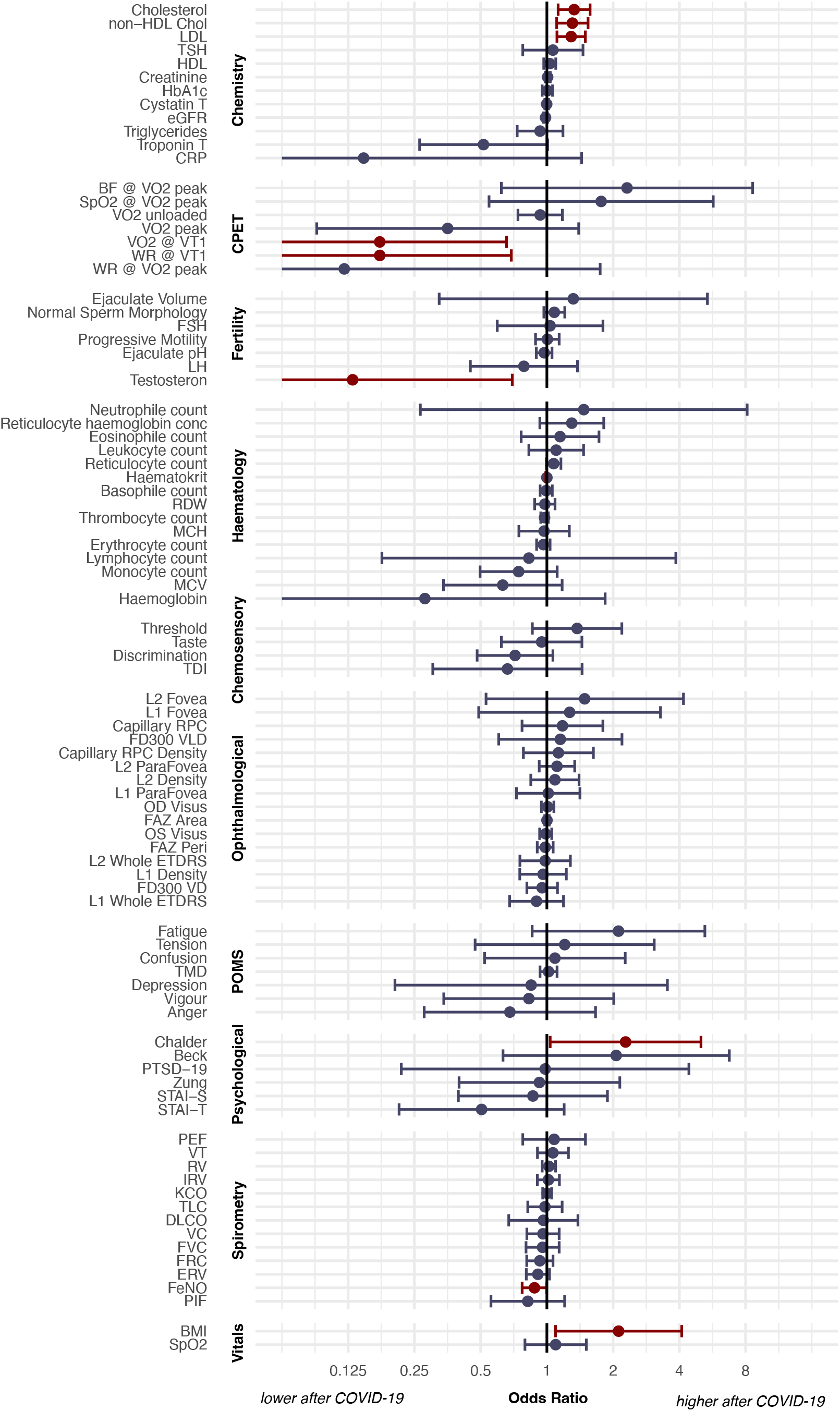
Multi-system sequelae after COVID-19. Volunteers more than 180 days after confirmed COVID-19 were compared to non-infected controls. Odds ratios for various parameters were calculated and are shown graphically with a 95% confidence interval. Values in red are significantly different after COVID-19 as compared to controls. We observed higher Cholesterol, non-HDL cholesterol, LDL, Body Mass Index and Fatigue (Chalder fatigue scale) after COVID-19, a lower oxygen uptake at the aerobic threshold (VO2 @ VT1) as well as a lower work rate at the aerobic threshold (WR @ VT1) and a lower testosterone level. Details for the values shown in this Figure can be found in Table 2.

**Figure 3:**
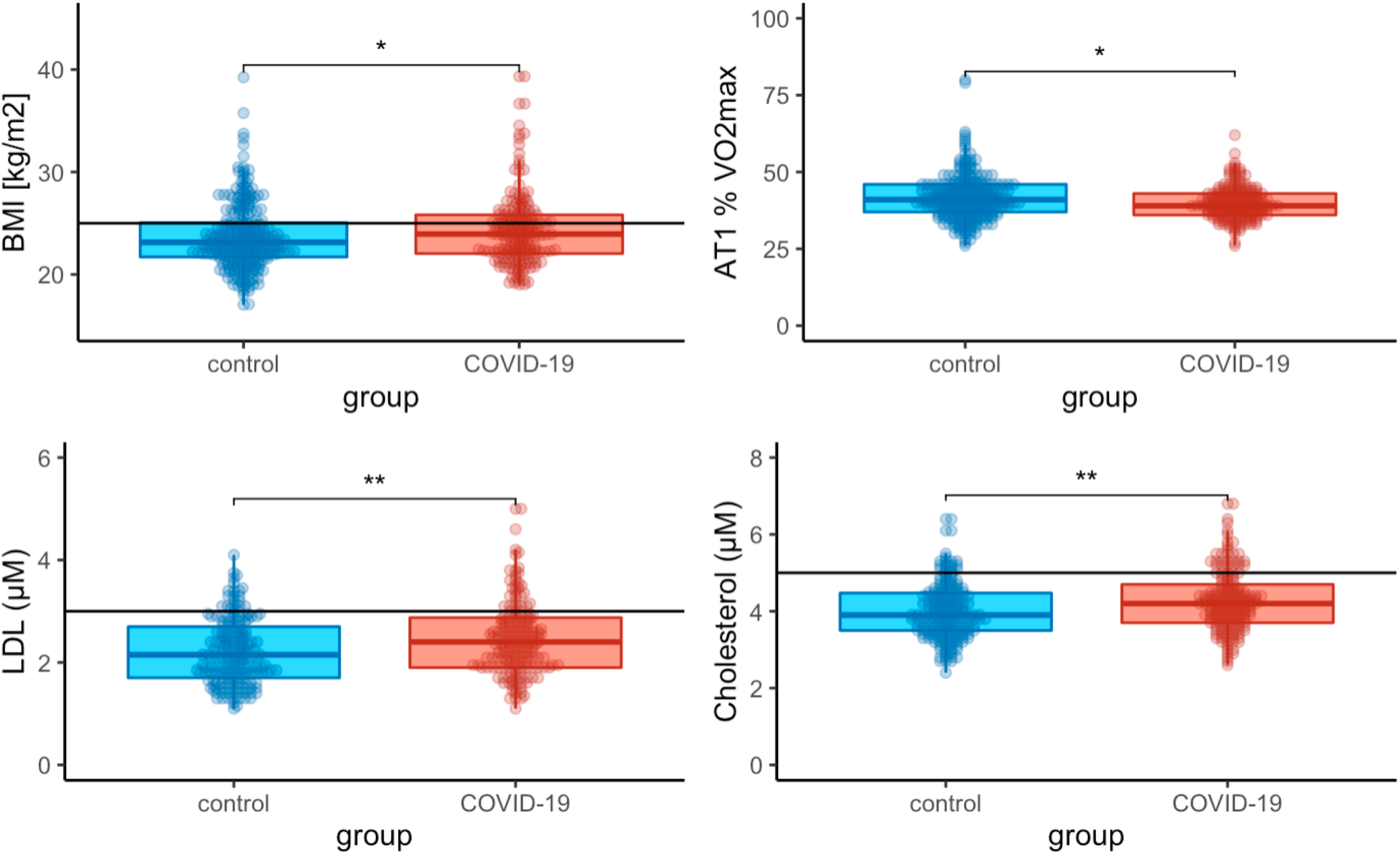
Signs of metabolic disorders after COVID-19. We observed significantly higher Body Mass Index (upper left), lower aerobic threshold (upper right), higher low-density lipoprotein (lower left) and higher cholesterol levels in patients more than 180 days after COVID-19 (red) when compared to non-infected controls (blue). Limits of normal values (25 kg/m2 for BMI, 5µM for total cholesterol and 3µM for LDL) are indicated as horizontal lines. * p<0.05, ** p<0.001

**Figure 4:**
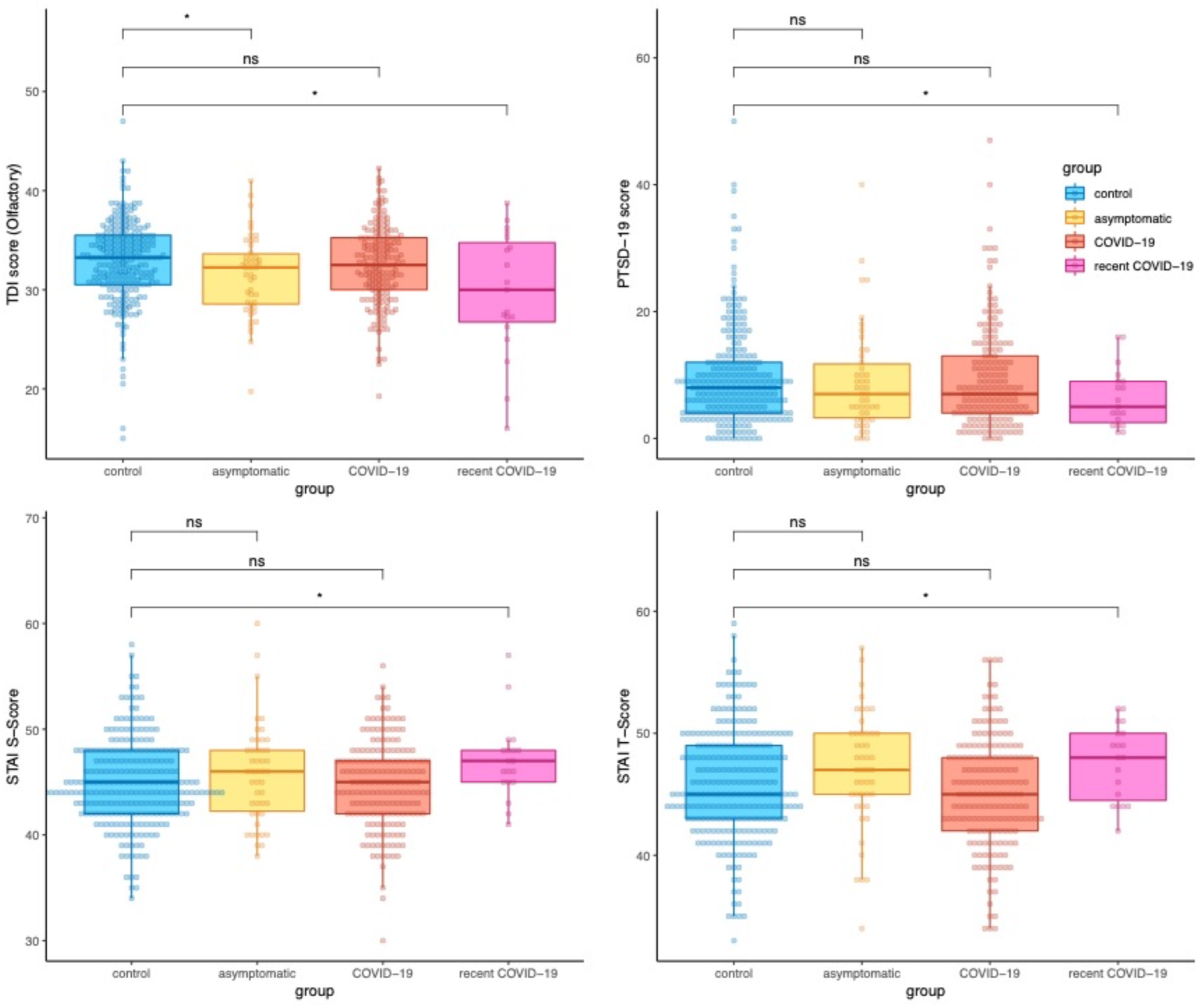
Parameters with significant change shortly after COVID-19, but not more than 180 days after COVID-19. We observed a significantly lower TDI (olfactory test) <180 days after COVID-19 (pink) but not >180 days after COVID-19 (red), indicating reversibility of hypo-/anosmia. In addition, Anxiety (STAI-S and STAI-T) as well as post-traumatic stress (PTSD-19) was different shortly after COVID-19 but this reversed after more than 180days back to levels comparable to controls. * p<0.05, ns=not significant

Unvaccinated, recovered, participants showed a limited capacity to neutralize SARS-CoV-2 regardless of the severity of the course of the previous infection. Vaccinated, recovered individuals exhibited a high titer of nAb, regardless of the severity of the course of the previous infection. Titers of vaccinated individuals were10-fold higher than those of recovered individuals.

**Table 2:**
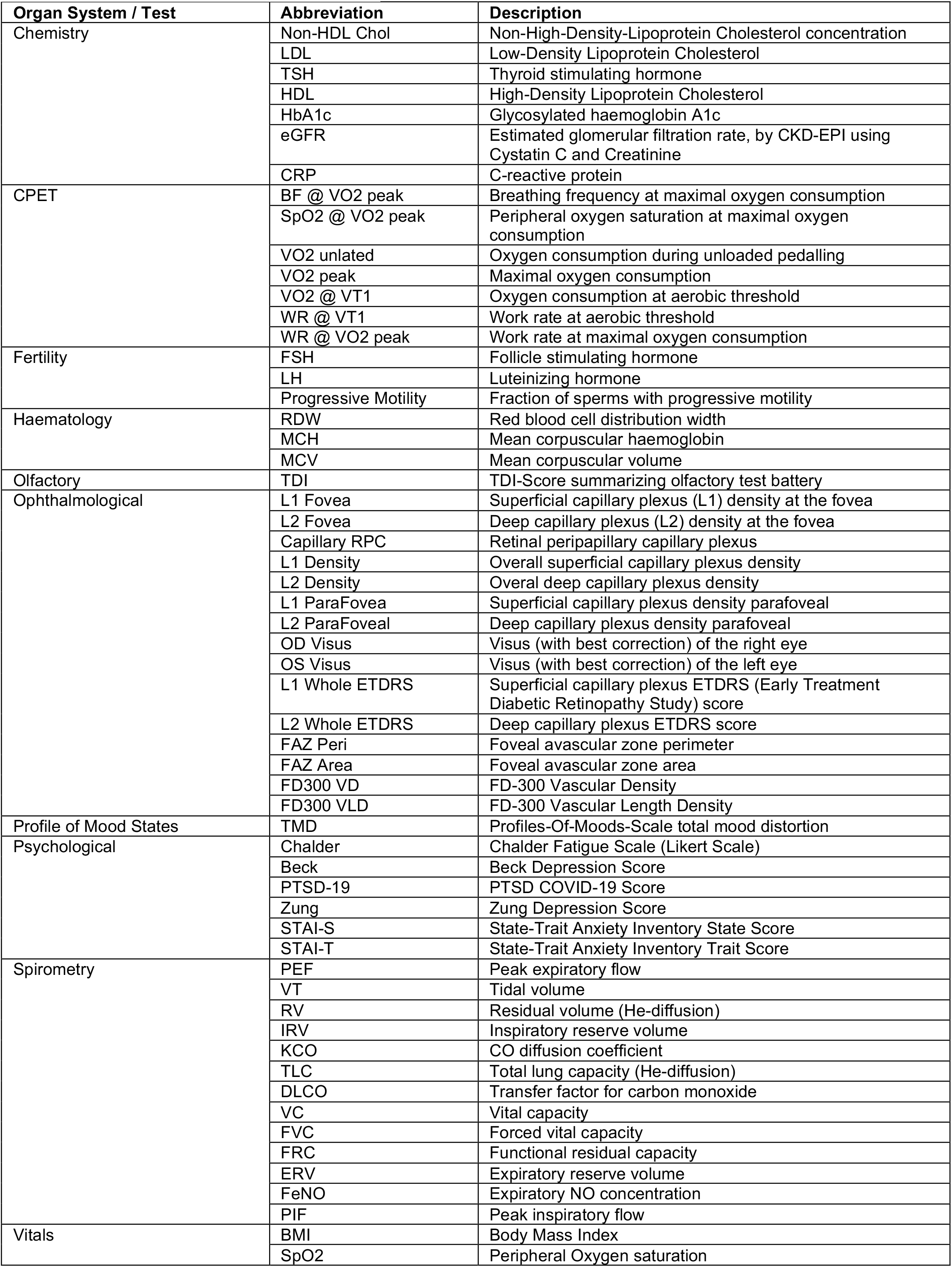
Legend for Figure 2

## Discussion

This comprehensive test series (evaluating cardio-vascular, pulmonary, neurological, ophthalmological, male fertility, psychological and general systems), administered more than 6 months after COVID-19 infection, showed significant sequelae; higher body mass index, dyslipidemia, and lower physical endurance. Such a constellation suggests that previously healthy young adults may have a higher risk of developing metabolic disorders and possible cardiovascular complications. Otherwise, the results of these quantitative analyses show overall recovery from mild COVID-19 and resolution of most sequelae at a mean of > 10 months post-infection. To date, this is the most comprehensive, controlled study, with the longest follow-up of sequelae in young, previously healthy adults. The multi-system impact of mild COVID-19 in this cohort with a mean age of 21 years,appears to be far less than that seen in older, multi-morbid or hospitalized patients. Overall, this is a positive perspective for young adult populations globally who have been infected with SARS-CoV-2. Regarding male fertility, it has been postulated that a SARS-CoV-2 infection may have potentially detrimental impact (20). In our subgroup analyses, we found evidence that recent infections (< 6 months before testing) were associated with poorer motile sperm counts but that this was no longer significant for non-recent infections. Our findings are corroborated by other studies. Donders et al found sperm quality to be sub-optimal post COVID-19 disease with an estimated recovery time of 3 months (21). In addition, we found significant hyposomia (TDI <31) in those infected in the previous 6 months. Observational studies of SARS-CoV-2 infected persons also report high levels of hyposomia. We recently followed up on army personnel using an App to self-report symptoms and found that positive-tested persons had a significantly reduced “sense of smell” (OR 18.24; 95% CI: 4.23, 78.69; p=0.00) compared to non-infected and that the hypogeusia persisted for a mean of 6.4 weeks (22). In addition, we found STAI S scores of anxiety levels to be significantly higher in recent COVID-19 participants. In an earlier study, Mazza et al (23) used questionnaires to screen for psychiatric symptoms in 402 adults one month post COVID-19 infection. A significant proportion of the participants self-rated in the psychopathological range for post-traumatic stress disorder (PTSD) (28%), depression (31%), anxiety (42%) and insomnia (40%). In our study, there were no significant differences in psychosocial questionnaire results between controls and those who had been infected more than six months previously. We consider the sequelae persisting beyond 6 months to be particularly important especially the excess burden of metabolic disorders including the elevated low-density lipoprotein and elevated total cholesterol. Our study could not differentiate whether COVID-19 in young adults predisposes for metabolic disorders or whether this predisposition existed previously and was accentuated by the infection. An earlier evaluation of the US Department of Veteran Affairs national healthcare database (6) found a substantial burden of health loss including diagnoses, medication use and laboratory abnormalities in patients with COVID-19 who survived at least for 30 days after diagnosis. The sequelae risk gradient increased according to the severity of the acute COVID-19 infection. Disorders of lipid metabolism were identified and an excess burden of use of antilipemic agents (6). Our findings also highlight lower physical endurance persisting many months post infection with significantly lower aerobic threshold (p=0.007). An earlier study of aerobic capacity in young Swiss army recruits (median age 21 years) (24), compared the results of physical endurance tests before infection to the same tests conducted 45 days post infection and found a significant decline in predicted maximal aerobic capacity in COVID-19 convalescent recruits. Our results suggest that this reduction in physical endurance can persist for longer than six months and we advocate further follow-up to define the duration of this sequela. Even mild infections with SARS-CoV-2 should not be underestimated (6,22,24). With the circulation of highly transmissible variants such as Omicron, a reduction in public health mitigation measures, a resumption of social activities (25), more and more young adults will have contact with SARS-CoV-2 and must live with consequences. Our study highlighted a positive sequela of COVID -19 showing that vaccinated, recovered individuals exhibited a high titer of nAb, regardless of the severity of the course of the previous infection. This further underpins the need to vaccinate persons recovered from COVID-19 regardless of the severity of their infection. The economic costs of even mild, long term COVID-19 sequelae and associated loss of productivity and possible need for disability allowances have still to be elucidated.

### Strengths and limitations

This is a unique cohort of young Swiss, mainly male, army recruits. In contrast to other studies, we had a control group, unequivocal evidence of SARS-CoV-2 infection and our test battery yielded objective and quantitative scores for analyses. A major strength of our study is the specifically designed, comprehensive test battery to quantify possible multi-organ sequelae based on the results of a systematic review (11).

A limitation of our study is the small proportion of female participants (5.6%) which precluded meaningful sex-based evaluation of sequelae in young women.

### Further research

The test battery developed here can be applied and even expanded for use in other population groups especially young women. We advocate further follow-up of this LoCoMo cohort and the initiation of other longitudinal studies to understand the trajectory of sequelae persistence beyond 1 year and focused research to clarify the pathophysiology and triggers of the identified sequelae and possibly the increased risk of cardiovascular disease. A clearer definition of Long-COVID or Post Acute Sequelae of COVID-19 (PASC) is also urgently needed (27) and we suggest that this should be nuanced for different population groups.

## Conclusions

Young, previously healthy, non-hospitalized individuals largely recover from mild infection and the multi-system impact of COVID-19 is less that seen in older, polymorbid or hospitalized patients. These results may be extrapolated to health-care workers and other young workforce adults and augur well for recovery in many body systems. However, as shown here, and in other studies, even mild infections in young adults can lead to sequelae that persist several months post infection with significantly more fatigue, hyposomia, poorer psychological scores and a short-term, negative impact on male fertility. Moreover, this controlled, cohort study with a long follow-up provided evidence of a constellation of higher body mass index, dyslipidemia and lower physical endurance even ten months post COVID-19 which is suggestive of a higher risk of developing metabolic disorders and possible cardiopulmonary complications. These results have societal and public health impact and can guide strategies for broad interdisciplinary evaluation of COVID-19 sequelae, their management, curative treatments, and support in young adult populations.

## Data Availability

The data produced in the present study may be available upon reasonable request to the authors

## Declaration of interests

We declare no competing interests.

## Contributors

PS and JWD designed the study and have access to all the data and take responsibility for the integrity of the data. PS, EL, TL, SZ, MIM, RZ, JWD contributed to data collation, NG and AS were instrumental in initiating the study, in financing and in liaising at the army/university interface. JWD did the data analysis. PS drafted the paper. All authors contributed to the revisions of the paper and to the approval of the final manuscript.

## Acknowledgements

We would like to thank all the SAF volunteers who took part in the LoCoMo study and who willingly gave their time, bio samples and data. We thank Dr. Christian Schmied for CPET instruction and Prof. Dr. Zeno Stanga for his constructive input to the paper. The following persons contributed in some way to study procedures: Anahita Bajka, Michel Bielecki, Martin Bosshard, Katja Bracher, Alon Cohen, Osman Efe Yoztekin, Lukas Egli, Rick Ernst, Anne-Sophie Ettlin, Nastasia Foa, Sara Fraefel, Kaylen Gähwiler, Susy Gutknecht, Michael Alexander Junker, Carola Kälin, Hatem Khrouf, Brigitte Leeners, Daniel Llanas Cornejo, Nico Marini, Raffaela Pitzurra, Isabelle Possa, Cécile Rasi, Manuela Rasi, Magdalena Rejdak, Livia Rentsch, Patricia Ritter, Adrian Rrhamani, Sadiq Said, Geraldine Schindler, Christina Schuler, Sophia Sidhu, Hanna Soffner, Shaymaa Soliman, Alexandra Veloudios, Sebastian Wyss and Xie Min.

Appreciation is due to Schiller Reomed AG and REAVITA AG for reduced rental fees for equipment used in our test battery.

## Notes

### Competing Interest Statement

The authors have declared no competing interest.

### Author Declarations

This study was approved by the Swiss Zurich Cantonal Ethics Committee (BASEC-Nr. 2021-00256).

